# Necessary for seizure forecasting outcome metrics: seizure frequency and benchmark model

**DOI:** 10.1101/2024.05.15.24307446

**Authors:** Chi-Yuan Chang, Boyu Zhang, Robert Moss, Rosalind Picard, M. Brandon Westover, Daniel Goldenholz

**Author notes:** **Corresponding author:** Daniel Goldenholz 330 Brookline Ave, Baker 5, Boston MA 02215. **Ethical statement** We confirm that we have read the Journal’s position on issues involved in ethical publication and affirm that this report is consistent with those guidelines. **Data Availability** Private data from Seizure Tracker and Empatica were made available upon request from these companies. These data are not public and may be requested by interested investigators subject to project approval. Source code is freely available here. https://github.com/GoldenholzLab/Metric_comparison_and_benchmark.git. **Ethics approval statement:** This study was deemed IRB Exempt by the BIDMC IRB.

## Abstract

Work is ongoing to advance seizure forecasting, but the performance metrics used to evaluate model effectiveness can sometimes lead to misleading outcomes. For example, some metrics improve when tested on patients with a particular range of seizure frequencies (SF). This study illustrates the connection between SF and metrics. Additionally, we compared benchmarks for testing performance: a moving average (MA) or the commonly used permutation benchmark. Three data sets were used for the evaluations: (1) Self-reported seizure diaries of 3,994 Seizure Tracker patients; (2) Automatically detected (and sometimes manually reported or edited) generalized tonic-clonic seizures from 2,350 Empatica Embrace 2 and Mate App seizure diary users, and (3) Simulated datasets with varying SFs. Metrics of calibration and discrimination were computed for each dataset, comparing MA and permutation performance across SF values. Most metrics were found to depend on SF. The MA model outperformed or matched the permutation model in all cases. The findings highlight SF’s role in seizure forecasting accuracy and the MA model’s suitability as a benchmark. This underscores the need for considering patient SF in forecasting studies and suggests the MA model may provide a better standard for evaluating future seizure forecasting models.

## Introduction

Many studies attempt to forecast seizures.^1–9^ However, patients’ seizure frequency (SF) is usually ignored when reporting the model performance. It has been observed across studies that patients have vastly different SFs.^10^ It is possible that model performance metrics calculated over a cohort might be influenced by SF and, thus, confound the evaluation of model performance.

Benchmark model selection is another important consideration. Often model performances are compared against a benchmark using random permutations (shuffling) of the predicted seizure labels (details below)^1,11,12^ Using permutation testing to assess a forecasting model is a very low bar to overcome, and probably does not have any clinical significance. Conversely, a moving average model (“what happened before is likely to happen again’) may be a better litmus test for a successful forecasting tool^12^.

We hypothesized that (1) there is a SF dependence that affects the performance of some forecasting metrics, and (2) using a moving average model is a better benchmark model compared to permutation testing. This study aims to explore these two hypotheses with simulation data and with two sets of real-world data.

## Materials and Methods

### Datasets and data preprocessing

#### Simulated dataset

We produced a structured simulation of 9 seizure diaries with 9 different seizure frequencies respectively. Each diary was a 10000-days-long binary array where 0 indicates there is no seizures and 1 indicates there is at least 1 seizure in that day. The monthly SF is determined by the number of seizure days in a month ranging from 1 seizure day to 9 seizure days per month. Of note, most patients from both clinical datasets had SF values within 1-9/month. All the seizure days occurred consecutively at the beginning of each month (Appendix). This organization of when seizures occurred was arbitrary – the key was that each diary had a prespecified number of seizures per month.

#### Clinical datasets

Two clinical datasets were evaluated, both approved by BIDMC IRB with Exempt status. We received access to the e-diary data through a data use agreement with Seizure Tracker LLC, facilitated by the International Seizure Diary Consortium. Seizure Tracker^10^ provided de-identified self-reported diaries. We selected patients based on the recording period and the length of diary (Appendix).

Another dataset was recorded by Empatica’s FDA-cleared Embrace 2, a wearable device for generalized tonic-clonic seizure (GTCS) detection.^9^ The device has a companion diary app “Mate” which patients sometimes use to manually enter seizures or to delete events that were false alarms. De-identified wearable-derived seizure diaries were provided by Empatica for the purposes of this statistical analysis. We selected patients based on recording duration and on the reliability of their e-diary interactions (Appendix).

### Metrics of interest

We focused on 4 commonly used metrics for forecasting: two for calibration (Brier Score and calibration curve) and two for discrimination (area under curve of receiver-operating characteristics (AUCROC), and area under curve of precision-recall curve (AUCPR)).^1,2,4–8,11,13^

To summarize results across diaries, we categorized each diary into SF bins ranging from 1 seizure day/month to 9 seizure days/month with a 1-seizure day/month bin size and reported the average within each bin. For results on extremely high and low SF, please see Appendix. The number of diaries in each bin was normalized by the total number of diaries for visualization purposes.

### Benchmark model: Moving average model vs. permutation testing

Moving average model (MA) is a simple causal forecasting model^12^. It predicts the probability of having seizure events by calculating the rate of seizure-present intervals (here, 24-hour intervals) in the diary history using a lookback window. In this study, we used a 90-day window, during which most SFs would be empirically expected to be steady.^8,10^ Since MA is intuitive and requires minimal computation (could even be computed manually by a patient/caregiver), we consider MA a candidate benchmark model.

Permutation testing is a widely used benchmark in forecasting tasks.^1,11^ It permutes the model forecasts and calculates the metrics of interest. This process is then repeated (e.g., 1,000 times). The average metric across all permutations is typically reported.

Improvement over chance (IOC) is another way to quantify a model performance, as shown in Eq. (1).

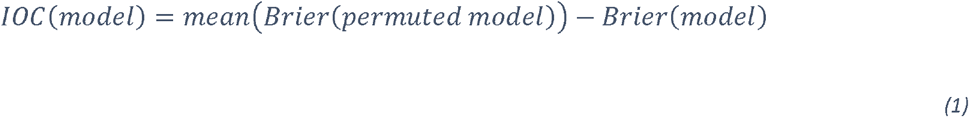

It can be shown that IOC for a perfectly accurate forecasting model (“truth”) is maximal (Appendix). Therefore, we consider the average result of permutations of truth, denoted permuted truth, as another candidate benchmark test, because it would provide the largest possible IOC for a given SF.

## Results

After preprocessing, there were 3,994 patients from Seizure Tracker with diary durations of 91-5,337 (median 525) days, and 2,350 patients from Empatica with diary durations of 90-1,551 (median 280) days.

Figure 1 shows the results of comparing the MA and permuted truth, across the four metrics for each of the three datasets. Seizure Tracker and Empatica have more diaries with low SF, as expected ^10,11^. In all twelve comparisons, the MA outperforms the permuted truth, showing MA is a harder baseline to beat. In nine of the comparisons the results depend on SF, usually improving with higher SF except in the case of the Brier Score, where lower (better) Brier score occurs with lower SF. The calibration curves of MA show slight overestimates in probability for low SF but improve as SF increases. All AUCROC values fluctuate around 0.5-0.6 across SF.

**Figure 1.**
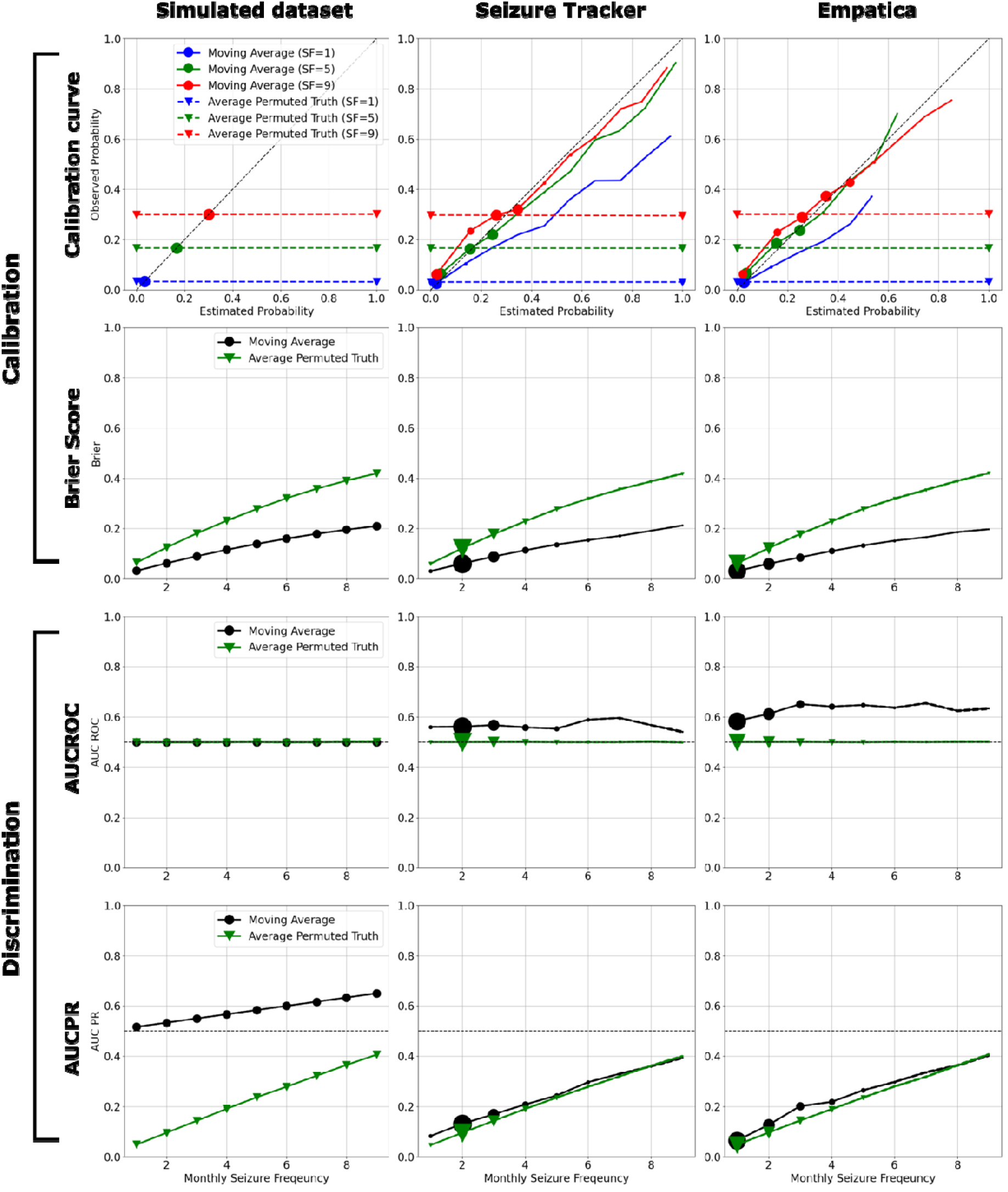
Twelve scenarios comparing the performance of MA vs. permuted truth. The calibration curve, brier score, AUCROC, and AUCPR are shown in rows. The results of simulated, Seizure Tracker, and Empatica datasets are shown in columns. In the calibration curves (first row), the monthly seizure frequencies 1, 5, and 9 are shown in blue, green, and red. The results of MA and permuted truth are indicated by solid line and dash line. The marker size indicates the normalized number of diaries within each estimated probability bin. Since the MA outcomes for the simulated dataset are constant, there is only one estimated probability in the calibration curve, resulting in a single marker instead of a solid line. The brier score, AUCROC, and AUCPR of MA and permuted truth are indicated by black and green solid lines respectively in the second, third, and fourth rows. The marker size indicates the normalized number of diaries within each SF bin. Note that in all twelve comparisons, MA performs as well or better than permuted truth. Additionally, within this range of SF, all metrics except AUCROC vary monotonically with seizure frequency. Higher SF values were explored in simulation (Appendix).

## Discussion

There are two main findings in our study. First, MA appears to be a better benchmark compared to permutation testing. Second, three of the metrics, the calibration curve, Brier score and AUCPR, show a dependence on SF while AUCROC appears to be relatively SF-independent.

Hence, it is necessary to report individual patient SF with these metrics when comparing model performances across different studies. Bins of seizure frequencies can be used if narrowly defined (as done here). When comparing models evaluated on datasets with different SF ranges, we suggest imputing metric performance for a common SF range and including SF independent metrics, such as AUCROC (Appendix). Critically, some SF values may not be very important to forecast (e.g., daily risk in patients who have a seizure per 2-days, or daily risk in patients with yearly seizures, etc.).

When comparing the performance of MA and permuted truth, we found MA always performs the same or better than permuted truth. Additionally, MA is preferable as a benchmark because it (1) is causal (i.e. does not require knowledge of the future), (2) is easily computed (“back of the envelope calculation”), and (3) is interpretable (“the previous seizure rate will recur”). Conversely, permutation is noncausal (knowledge of the future is required) and requires more computational resources. Anecdotally, our investigations have found MA to be surprisingly accurate in multiple seizure forecasting contexts and therefore a more challenging benchmark to overcome for a candidate model.

There were some differences between the simulation and the clinical datasets. Many of these differences reflect the simplistic assumptions used for the simulation, as well as methodological choices made for our study (Appendix).

The emphasis of this paper is identifying mathematical guideposts for testing algorithms. In contrast, the *value* of seizure forecasting tools is beyond the scope of this study; patient attitudes, beliefs, desires, and behaviors all need to be accounted for prior to deploying a forecasting tool.

In summary, this study provides insight into the importance of including patients’ seizure frequency in seizure forecasting tasks and demonstrates that MA represents a valuable benchmark with minimal computational complexity.

## Data Availability

Private data from Seizure Tracker and Empatica were made available upon request from these companies. These data are not public and may be requested by interested investigators subject to project approval. Source code is freely available here. https://github.com/GoldenholzLab/Metric_comparison_and_benchmark.git

## Acknowledgements

Thanks to the International Seizure Diary Consortium for facilitating data sharing. DG and CC are supported by NINDS K23NS124656. BZ is supported by T32 HL007901-25. Dr. Westover was supported by grants from the NIH (R01NS102190, R01NS102574, R01NS107291, RF1AG064312, RF1NS120947, R01AG073410, R01HL161253, R01NS126282, R01AG073598), and NSF (2014431).

## Author contributions

CYC – drafting, editing, data analysis, data interpretation

BZ – data acquisition, data analysis, editing, data interpretation

RM – data acquisition, editing, data interpretation

RP – editing, data interpretation

MBW – editing, data interpretation

DMG – conception and design, editing, data interpretation

