## Appendix for "Necessary for seizure forecasting outcome metrics: seizure frequency and benchmark model"

### Dataset

#### Simulated dataset

An illustration of simulated diaries is shown. The red section indicates the seizure days.

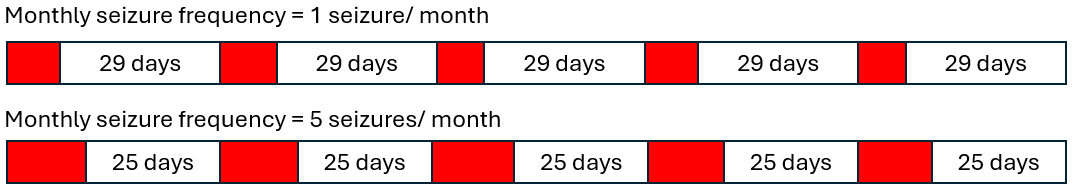

*Supplement Figure 1. Illustration of simulated diaries.*

There is no special reason for organizing the diary like this rather than randomly spreading the seizures through the month in a permuted fashion. The exact order is not important, as long as a specific seizure rate is respected. We selected this organization because it was simple.

#### Seizure Tracker dataset

The Seizure Tracker was determined exempted by the Beth Israel Deaconess Medical Center Institutional Review Board. Demographic and other characteristics of this data has been previously reported by Ferastraoaru et al 2018.

The original dataset contains diaries from 21,318 patients. We first selected patients with recording date started after December 1^st^, 2007, and ended before August 8^th^, 2022. Secondly, we include patients with diary length ≥ 90 days to ensure robust SF estimation. We estimated SF by calculating the mean seizure days over the entire diary and removed patients with estimated SF in [0.5,9.5], to remove outliers.

#### Empatica dataset

De-identified data from Empatica Embrace 2 was obtained in accordance with the user agreement/privacy policy. All Empatica data analysis was performed by Empatica employees (BZ, RP) to ensure these de-identified data were handled with additional safeguards for privacy.

An FDA-cleared machine learning algorithm on the smart wristband detects generalized-tonic-clonic seizure GTCS in real-time and stores the timestamped alert. Patients can cancel a detection as a false positive during alert onset. They can also revise the alert records later, including labeling false positives, adding false negatives (missed detection), and confirming true positives. Users who did not consistently report false positives were excluded from further analyses. All GTCS alert records from the wristband are de-identified.

Excluding low-quality raw data, 80,424 GTCS (median=34, range 3-373) were recorded from 2,350 patients. The median length of longitudinal GTCS records was 280 days (range 90-1551). 50.5% (N=1,187/2,350) of the patients were male, and 48.3% (N=1,135/2,350) were female. 1.2% (N=28/2,350) of the patients reported other genders. At the beginning of the records, 63.8% (N=1,499/2,350) of the patients were adults (18 and older), and 36.2% (N=851/2,350) were children. The subtype of epilepsy was not available.

### Proof of maximal IOC

Statement:

$$IOC\left( Model \right) \leq IOC\left( Truth \right)$$

That is:

$$mean\left( Brier\left( permuted model \right) \right)-Brier\left( model \right)\leq mean\left( Brier\left( permuted truth \right) \right)-Brier\left( Truth \right)$$

Let:

1. $Y_{m}$ = Model forecast
2. $Y_{t}$ = Ground truth
3. $Y_{mp}$ = Permuted model forecast
4. $Y_{tp}$ = Permuted ground truth
5. $n$= Number of samples
6. $m$= Number of positive samples in ground truth
7. $q$= Number of positive samples in model

The statement can be rewritten as:

$$mean\left( Brier\left( Y_{mp} \right) \right)-Brier\left( Y_{m} \right)\leq mean\left( Brier\left( Y_{tp} \right) \right)-Brier\left( Y_{t} \right)$$

At position $i,j$:

(a) $y_{i},y_{j}$ = True labels where $y_{i}\neq y_{j}$
(b) $\hat{y_{i}},\hat{y_{j}}$ = Model forecasts
(c) $Brier\left( Y_{m} \right)=\frac{1}{n}\left( \left( y_{i}-\hat{y_{i}} \right)^{2}+\left( y_{j}-\hat{y_{j}} \right)^{2}+\sum_{k,k\neq i,j}^{n} \left( y_{k}-\hat{y_{k}} \right)^{2} \right)$
(d) After swapping model’s outcome at $i,j$:

$Brier\left( Y_{m},swap i,j \right)=\frac{1}{n}\left( \left( y_{i}-\hat{y_{j}} \right)^{2}+\left( y_{j}-\hat{y_{i}} \right)^{2}+\sum_{k,k\neq i,j}^{n} \left( y_{k}-\hat{y_{k}} \right)^{2} \right)$

(e) $Brier\left( Y_{m},swap i,j \right)-Brier\left( Y_{m} \right)=$

$$\frac{1}{n}\left( \left( y_{i}-\hat{y_{j}} \right)^{2}+\left( y_{j}-\hat{y_{i}} \right)^{2} \right)-\frac{1}{n}\left( \left( y_{i}-\hat{y_{i}} \right)^{2}+\left( y_{j}-\hat{y_{j}} \right)^{2} \right)$$

$$=\frac{1}{n}\left( -2y_{i}\hat{y_{j}}-2y_{j}\hat{y_{i}}+2y_{i}\hat{y_{i}}+2y_{j}\hat{y_{j}} \right)$$

$$=\frac{2}{n}\left( -y_{i}\hat{y_{j}}-y_{j}\hat{y_{i}}+y_{i}\hat{y_{i}}+y_{j}\hat{y_{j}} \right)$$

(f) When $\left( y_{i},y_{j} \right)=\left( 0,1 \right)$ : $Brier\left( Y_{m},swap i,j \right)-Brier\left( Y_{m} \right)=\frac{2}{n}\left( \hat{y_{j}}-\hat{y_{i}} \right)$
(g) When $\left( y_{i},y_{j} \right)=\left( 1,0 \right)$ :$Brier\left( Y_{m},swap i,j \right)-Brier\left( Y_{m} \right)=\frac{2}{n}\left( \hat{y_{i}}-\hat{y_{j}} \right)$
(h) The maximal (f) and (g) happen when:

$\left( \hat{y_{i}},\hat{y_{j}} \right)=\left( y_{i},y_{j} \right)$ :$\max\left( Brier\left( Y_{m},swap i,j \right)-Brier\left( Y_{m} \right) \right)=\frac{2}{n}$

(i) To maximize $mean\left( Brier\left( Y_{mp} \right) \right)-Brier\left( Y_{m} \right)$, all the swaps should be maximized. Thus, $Y_{m}$ must be binary.
(j) According to (a) and (h), number of valid swaps ($y_{i}\neq y_{j}$) for $Y_{tp}$ is always greater or equal to number of valid swaps ($y_{i}\neq y_{j} and \hat{y_{i}}\neq\hat{y_{j}}$) for any binary model $Y_{mp}$
(k) To maximize number of valid swaps for $Y_{mp}$, the set ($y_{i}\neq y_{j}$) must be equal to the set ($y_{i}\neq y_{j} and \hat{y_{i}}\neq\hat{y_{j}}$), that is, $q=m$
(l) According to (i) and (k), $\max\left( mean\left( Brier\left( Y_{mp} \right) \right)-Brier\left( Y_{m} \right) \right)$ happens when $Y_{m}=Y_{t}$

Therefore:

$$IOC\left( model \right)\leq IOC\left( Truth \right)$$

Thus, permuted truth versus true forecast will provide the highest value of IOC compared to IOC using the permuted version of any other forecast with the same ground truth.

### Difference between the simulation and real-world data

The MA in the simulated dataset only has a constant outcome because the 90-day window perfectly covered 3 seizure cycles. Hence, its calibration curve only has one estimated and one observed probability for each SF. In the clinical datasets, there is an overestimation in the calibration curves of MA because MA equally spreads out the probability of having a seizure day within 90-day window. This contamination is more severe at low SF because there are more non-seizure days within the 90-day window. Brier score is sensitive to imbalanced dataset. A model which performs better in the majority of situations has a better (lower) Brier score. For both permuted model and MA, the expectation of model outcome is a constant, SF. In the case of constant expectation, the Brier score will become higher (i.e. lower accuracy) with increases class balance and reach the maximum when the classes are completely balanced (50/50).

The AUCROC of permuted truth is very close to 0.5 across SF for each dataset because the expected value of permuted truth outcome is SF, causing its ROC to be nearly diagonal. The same phenomenon happens in AUCROC of MA in the simulation. On the other hand, though the expected value of MA in the clinical datasets is also SF, MA has AUCROC slightly above 0.5 due to its ability to capture slow temporal fluctuations in diaries. The AUCPR of both permuted truth and MA increase with SF because both models have poor discriminative ability, causing the precision to be very close to SF when recall equals 1. Of note, the AUCPR of MA in the simulation performs differently compared to other datasets. Because the outcome of MA is a constant in the simulation, its curve of precision and recall are nearly off diagonal with the precision equals to SF when recall equals to 1. The AUCPR of MA is lower in the clinical datasets than in the simulated dataset because there are more outcome values from MA and thus generating more precision data points as recall goes up. The more data points, the more accurate the precision-recall curve is. An example of Precision and Recall curves of MA from the bin with monthly SF=2. Each line is one diary. We can see most of the precision drops rapidly as recall increases in Seizure Tracker.

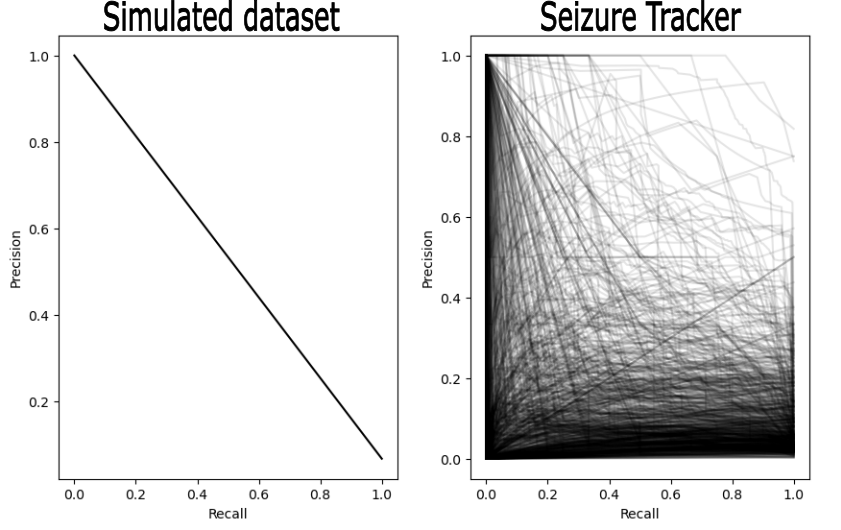

*Supplement Figure 2. An example of Precision and Recall curves of MA from the bin with monthly SF=2. Each line is one diary.*

### Example: Comparing models from different datasets

Model A was evaluated on a dataset with 2≤monthly SF≤4 and got a Brier score = 0.15. Model B was evaluated on another dataset with 6≤monthly SF≤8 and got a Brier score = 0.18. Though model A has a Brier score lower than model B, one cannot claim model A performs better than model B since model A is evaluated on a dataset with lower SF range. According to Figure 1, Model A is worse than MA while Model B is better than MA at given SF ranges. One possible way to handle different SF ranges is to interpretate model’s metric to the same SF range using imputation methods. Alternatively, model A and model B should be compared within the same SF ranges.

### Limitation of the study

There are some limitations in this study. First, it is impractical to evaluate all the metrics used in forecasting tasks. Therefore, our findings might not be generalizable for certain metrics. Nevertheless, we examined commonly used metrics and suspect other metrics will have similar findings. Secondly, the 90-day window used to estimate SF in this study was selected empirically. Although 90-day was appropriate for the SFs and models examined here, other window sizes may be appropriate for certain SFs or models. Lastly, our clinical data didn’t fully explore the seizure frequency dependency for extremely low or high SF. See section on “Full range of SF”.

### Using different window sizes

This study used a 90-day window for the moving average (MA) model to look back and determine the typical seizure rate. The number was selected empirically based on the typical seizure rates noted in both clinical datasets such that most typically reported seizure rates would be captured. However, there are circumstances when a 90-day window would not be helpful. For example, if a patient has a typical seizure rate of yearly, or even 2 per year, a 90-day window will mis-represent the daily risk. Similarly, if a patient has daily seizures, a 90-window would be functional, but a shorter window size would likely provide more accuracy for shorter term risk changes. Therefore, we recommend using a 90-day window in the case when seizure rates are unknown, but rather to adjust the lookback window in cases where a longer or shorter window would provide better insight into ongoing risk.

In a similar way, the 24-hour risk forecast discussed in this manuscript was a choice of convenience. It would be very helpful for patients who have infrequent seizures to have daily risk for seizures available. However, patients who have very frequent (i.e. higher than 15 per month) rates might prefer more granular forecasts, such as AM/PM, hourly, etc. Some algorithms or biomarkers would be more suitable to smaller or larger forecast horizons than precisely 24 hours as well. If one changes the forecast horizon duration to be larger or smaller, the basic ideas presented in this study are still relevant. Seizure frequency will still have an effect on performance metrics, and MA would still be recommended as a more appropriate benchmark compared with a permuted benchmark.

Several examples can illustrate both points. Suppose a patient has daily seizures lasting 30 seconds. Such a patient would benefit from a smaller window size from the moving average, perhaps even 1 week would be sufficient. At the same time, the forecasting horizon in that patient would be more useful on an hourly basis rather than daily. A second patient perhaps has a seizure once in 6 months. This patient would likely benefit from a much longer window, perhaps 2 years long, to make basic estimates of risk. The forecast window for this patient could be daily but might make more sense at a weekly level. In both patients, candidate forecasting tools can be tested at different SF values, and compared to an appropriate MA model.

### Seizure clusters

Seizure clusters occur in 25-50% of people with epilepsy. Sometimes these clusters may reflect statistical fluctuations, other times they might represent prolonged periods of elevated cyclical seizure risk, and other times they represent uniquely dangerous periods due to a combination of factors that conspire to temporarily increase risk much higher than usual. Regardless, our current analysis does not formally account for clusters. Classically defined clusters would be X seizures within 12 or 24 hours. Our main analysis focused on seizure days as a binary quantity, thus a cluster within a day would simply be counted as a seizure day. More broad definitions would include groups of seizures occurring with an unusually for that patient inter-seizure interval. In that latter case, some patients would be seen as having a cluster of seizure days. The proposed benchmark MA model would handle classical clusters as single events and cluster of days would be treated the same way un-clustered seizure days would be. This imples that MA is agnostic to the existence of multi-day clusters, and is blind to within day clusters, when the forecast horizon is 24 hours. Of course these facts would change for different choices of MA window size and forecast horizon sizes. We do not claim that MA is “smart” nor that it is well suited to clusters. Indeed, one could construct a simple forecasting tool that understands clusters, and for some patients that tool would beat MA. Because clusters are poorly defined and poorly understood, we recommend setting the benchmark to MA knowing that it has blindspots to clusters.

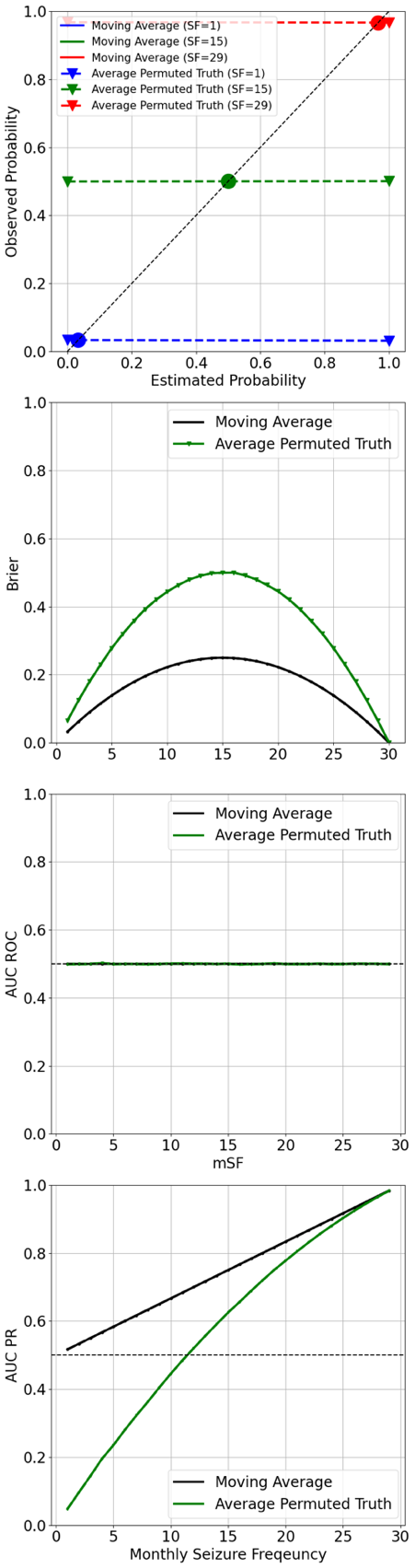

### Full range of SF

The results of full range SF (0≤SF≤30) on simulated dataset shown in Supplement Figure 3 indicates that MA performs better than permuted truth at all SF.

*Supplement Figure 3. The performance of MA and permuted truth across full range seizure frequencies in simulated diary data.*

### Clinical impact

Choosing a statistically meaningful, high performing seizure forecasting tool is only the first step in a larger set of challenges. The first hurdle is demonstrating that a tool is able to beat MA at the SF one is interested in. The second hurdle is: does the tool have a clinical impact? Would patients with that SF be interested in using this tool? Would it change their behavior in positive or negative ways? Would the patient get more harm or benefit from using this tool?

Even if clinical impact were assured, there are many ways to present the same information. Should it be auditory, visual, a combination, or user-selectable? Should the forecast provide a binary yes/no type of information based on mathematically or user-selected cutoffs, or should it provide raw probabilities? Should the tool indicate exactly how inaccurate it is whenever presenting new information, or should that description of inaccuracy be hidden from view unless called up? Can software or hardware that presents seizure forecasts be considered a medical device that should be subject to government regulation? Must it be? What are the minimum levels of accuracy and harm-reduction regulators should demand of these tools if they should be regulated? What are the minimum levels of accuracy that patients would prefer? If forecasts were imperfect, would it be allowable to use them as inputs to “closed loop” neuromodulatory devices? If yes, how imperfect is sufficiently safe?

The above questions are presented to highlight that the present work is focusing on an early question about mathematical value, which must be contextualized in a much larger set of questions that are medical, psychological, sociological and regulatory.
